# A Novel Approach for Estimating the Final Outcome of Global Diseases Like COVID-19

**DOI:** 10.1101/2020.07.03.20145672

**Authors:** Demetris T. Christopoulos

## Abstract

The existence of a universal law which maps the bell curve of daily cases to a sigmoid curve for cumulative ones is used for making robust estimations about the final outcome of a disease. Computations of real time effective reproduction rate are presented and its limited usefulness is derived. After using methods ESE & EDE we are able to find the inflection point of the cumulative curve under consideration and study its time evolution. Since mortality processes tend to follow a Gompertz distribution, we apply the properties of it and introduce novel estimations for both the time remaining after inflection time and the capacity of the curve. Special properties of sigmoid curves are used for assessing the quality of estimation and as indices for the cycle completion. Application is presented for COVID-19 evolution for most affected countries and the World.

## 1. Introduction

Modern Epidemiology is based merely on the mathematics of dynamical systems that describe the time evolution of basic epidemiological variables, after having established a set of causal relations between them. Although it is a well known mathematical field which has given many successes, the fact that those models need calibration for determining the value of parameters and their tendency to give exponential growth for the mid and long term output makes them not suitable for all diseases. Another disadvantage is the extensive uncertainty intervals given for each initial estimate that makes all predictions uncertain and closer to “alarm calls” rather than to scientific results. After COVID–19 our life has drastically changed due to the social distancing measures, thus we need reliable tools for estimating the time evolution and the final outcome per country.

Current work focus on the properties of every cumulative curve, either confirmed cases, deaths or recovered, in order to make robust inferences about both remaining time and final outcome at the end of current disease cycle. The only needed requirement is a small initial waiting time until an inflection point has occurred.

Structure of the paper: introduction, models & limitations, introducing alternative view, comparison of the two approaches, time dynamics of COVID–19, discussion.

## 2. Broadly Accepted Models, Limitations and Failures

### 2.1. Short Review

Modern mathematical epidemiological models can be classified in a few categories, the *compartment models*, [17], [18] with their enhancements, extensions, generalizations and the *stochastic models*, see [1] for an extensive presentation. A short history can be found in [3], the three basic models (*SIS, SIR, SIRS*) are being explained in [14] while from the mathematical point of view a rigorous classification is presented at [15] and a more detailed presentation with example codes can be found in [16]. The procedure followed by those dynamical systems is (i) the creation of causal differential or difference equations between the variables Susceptible (S), Exposed (E), Infected (I), Recovered (R), Deaths (D) or others model dependent ones, (ii) calibrating the model to real time data and estimating its parameters, (iii) using it for making predictions about the time evolution of the disease.

A typical epidemiological model can be described by the set of differential equations of Eq. 1, for approximately continuous time *t* > 0 

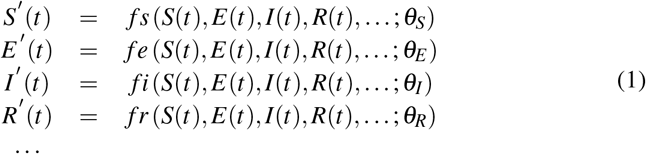

or by the set of difference equations of Eq. 2 for discrete time *n* = 0, 1, …, usually in days or weeks

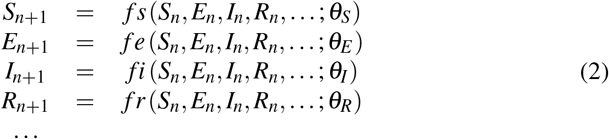

Here *S, E, I, R*,… stand for the fractions of Susceptible, Exposed, Infectious, Recovered,… respectively of the total population and *θ*_*S,E,I,R*,…_ is the relevant vector of parameters needed for modeling each variable. The functions that describe the rate of evolution can be either linear or non linear, while the parameters can be fixed or time dependent too, so an extended class of models can be generated by the above presented ‘equations of motion’.

As every typical useful dynamical system, there always exist an equilibrium state for the models of Eq. 1 which describes the long term behavior for the population, after the beginning of the disease. That process may or may not be oscillating, depending on the estimated value of the involved parameters.

Our task is not to present an extended literature review of epidemiological models, but to compare their predictions with the non parametric treatment of our work.

### 2.2. The Exponential Growth Does not Last for Long Time

One of the many slightly declared assumptions is that when a country is infected by a disease and if any kind of *Non Pharmaceutical Interventions* (*NPI*) will be applied, then both confirmed cases and deaths will continue to increase exponentially, following the initially calculated basic effective rate *R*_0_, so a very rough estimation for the expected confirmed cases after time *t* will be 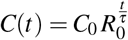, with *τ* the serial interval or generation time. For example if we take the case of UK and compute its *R*_*t*_, after using [10] with serial interval 4 days (*sd* = 4.75) & weekly smoothing, then from Fig. 1 we see that on March 14th *R*_*t*_ *≈* 2 while from [4] we have *C*_0_ = 1144, thus the exponential phase could be roughly described by the above simple formula. so the expected cases, given *τ* = 4 days, [20], will be after a month *C*(30) ∼ 207*K*, much greater than the reported value of 90K. If we recall that real number of infected cases is many times the reported ones, [2], then above estimation could further increase and give unacceptable large numbers. A desired feature for each model would be is its ability to predict the inflection point of the cumulative confirmed cases or deaths, since after that point situation tends to enter a decreasing mode, at least for the cycle under consideration.

**Figure 1:**
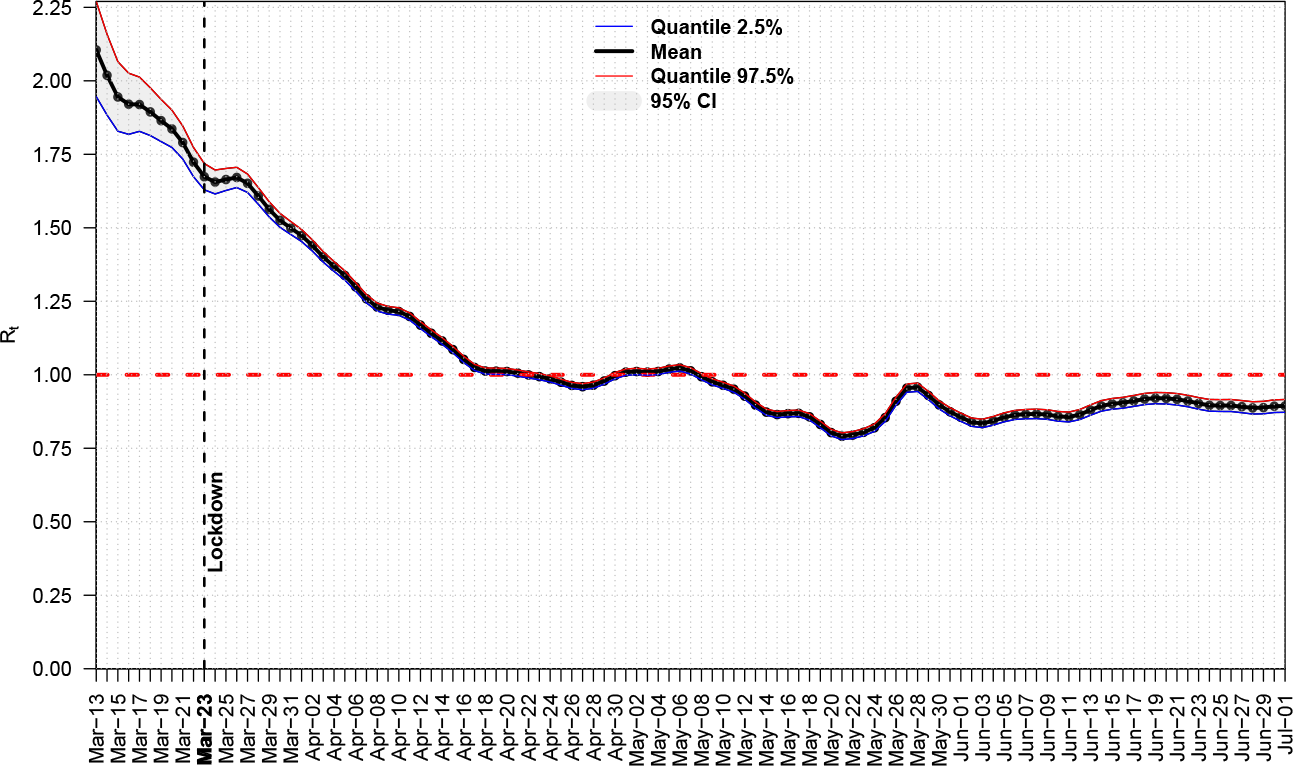
Real Time *R*_*t*_ for UK

### 2.3. Limitations of Effective Reproduction Rate R_t_

During recent COVID–19 pandemic most of the countries applied *NPI* in order to reduce the *R*_*t*_ below the unity, however even if they succeeded to keep it permanently there, no effective improvement was achieved, despite the commonly accept rule that a disease has been controlled that way. For example Italy after March 27 had *R*_*t*_ *<* 1, but that did not make any difference in both daily confirmed cases or deaths. The same holds for all heavily affected Western countries, Spain, France, UK, US have followed a similar pattern, national lockdown, keeping the rate below unity and the time evolution of the virus was not affected.

On the other hand there existed countries like Greece for which after releasing lockdown the daily computed *R*_*t*_ was many times above 1 and nothing bad was happened, see Fig. 2 (after using [10]), thus reproduction rate cannot serve as a reliable proxy for estimations, it can serve as just a picture of the currently observed disease spread dynamics.

**Figure 2:**
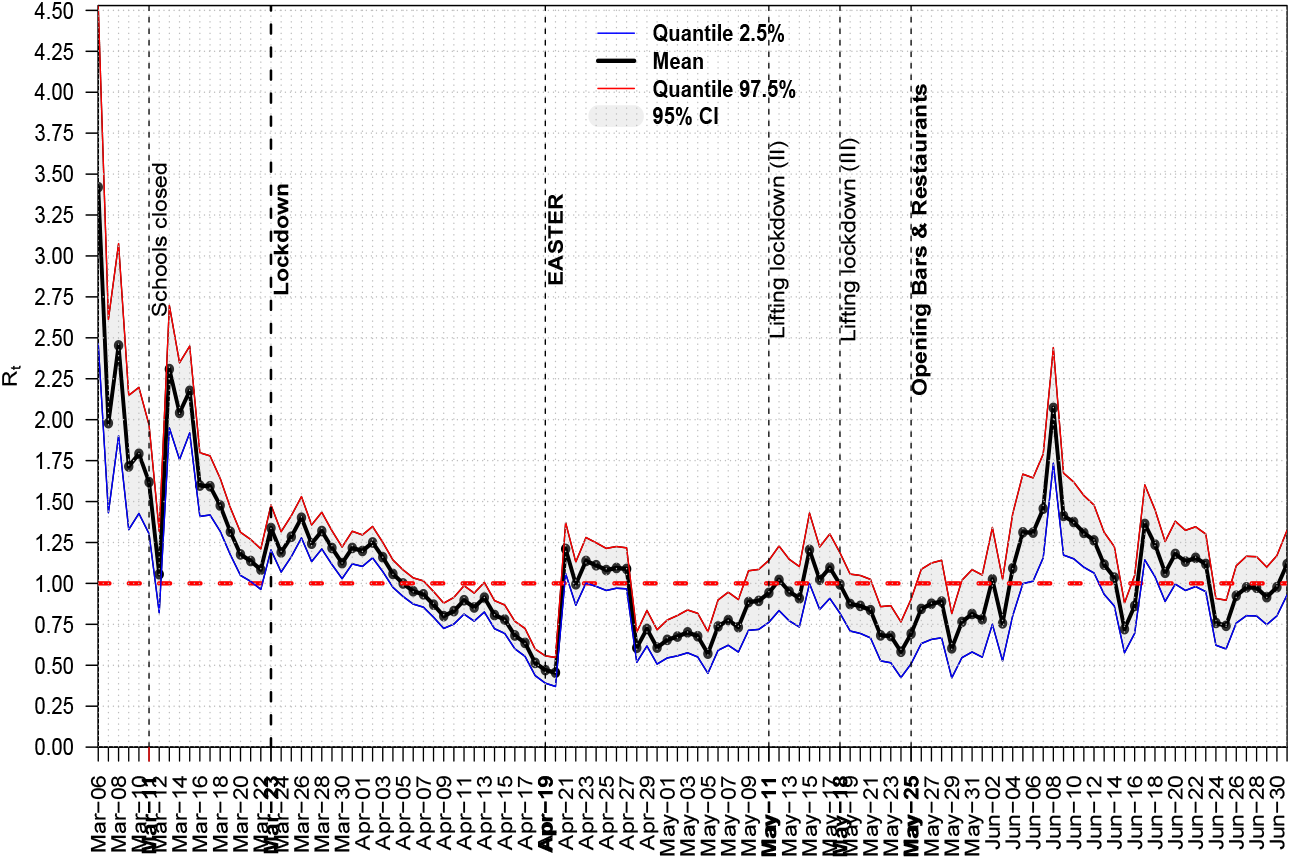
Daily *R*_*t*_ for COVID–19 in Greece

### 2.4. Failure of Model Predictions

Mathematical models can be calibrated and fitted to the data under a specific time period with success, see for example [12], however when a mid to long term forecasting is requested, then they are giving extremely large interval estimations with mean values also very large, compared to the actually realised a few months later.

For example at the beginning of COVID–19 in US & UK a study estimated the overall deaths at extremely large numbers, 510K & 2.2M respectively, just for a seven month window, see Fig. 1 of [11]. Such estimations rang as an alarm for taking social distancing measures in UK, however, even after adopting suggested interventions, the disease did not slowdown its evolution. Currently UK, US have 44K, 128K of deaths, much smaller than the predicted ones.

Even for a country like Greece, which was one of the countries that took measures very early and succeeded to combat the disease, a relevant study after using a *SEIR* model was predicting a cumulative number of confirmed cases approximately at 12K with a confidence interval [4.5K,27.5K], see Fig. 3 of [21]. Currently Greece has a cumulative number of 3.4K confirmed cases, smaller even from the lower edge of predicted interval.

**Figure 3:**
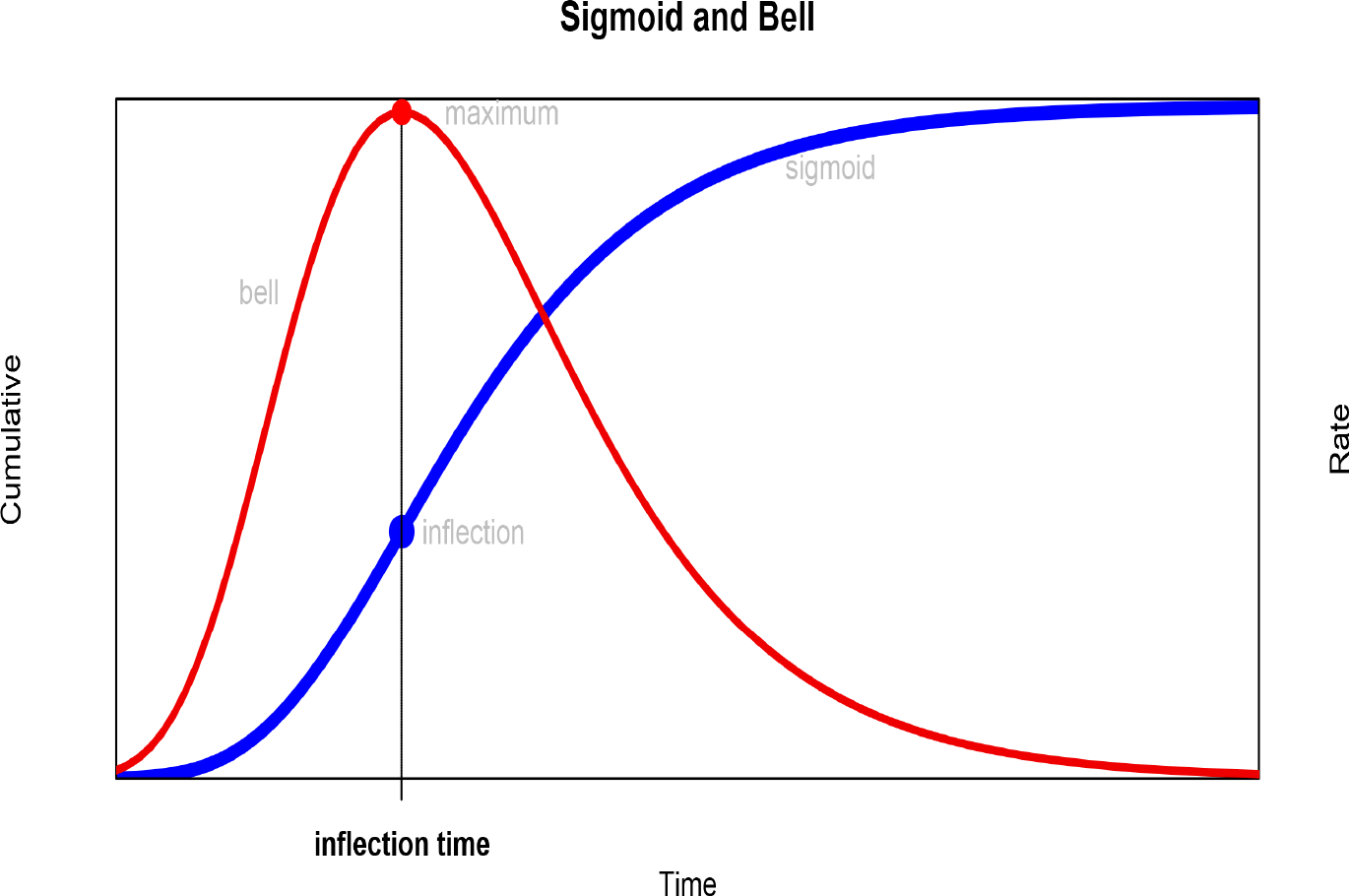
Bell and Sigmoid

## 3. The Alternative View: Non Parametric Inflection Analysis

### 3.1. Bell and Sigmoid Curve

Any disease can be considered as a time evolved process which has a daily output, either confirmed cases, deaths or recovered representing the rate while the relevant cumulative variables give the final outcome from the beginning and are always S-shape curves or sigmoids. That is a kind of universal law which determines all processes and maps the bell curve of rate to the sigmoid curve of total outcome, see Fig. 3 and observe that maximum of daily rate corresponds to the inflection of the cumulative outcome. More law examples for a broad range of processes can be found at [19]. Why do we care about inflection point so much? Well contrary to the rate curve, the sigmoid one is more smooth due to the relative small changes added every day. That works by itself as a kind of smoothing to the added noise and makes things more easy from a computational aspect.

Mathematically speaking Gompertz [13] curve which has been showed that describes mortality processes with its 1^*st*^ and 2^*nd*^ derivatives are defined at Eq. 3, 4, 5, *L, b, κ* > 0.

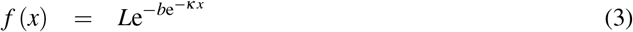

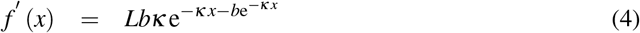

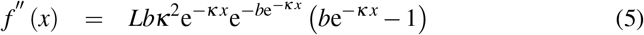

It is evident that inflection point is 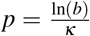 and we can check that corresponds to the maximum of the bell curve. Since the final outcome of the cumulative sigmoid curve is just *L*, we conclude that it holds Eq. 6 and that means we are able to make a first estimation for the capacity of the curve *L*: if we have a method to compute the inflection point *p*, then a first estimation for the expected capacity would be just *e* times the observed cumulative outcome at the time of inflection point.

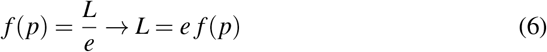

Thus we have proven next Lemma 3.1, or more conveniently the ‘*e–rule*’.

#### Lemma 3.1.

*If a cumulative variable like the total confirmed cases or deaths for a disease (i) resembles a Gompertz sigmoid pattern & (ii) has reached its inflection point, then its final maximum will be at least e times the observed value at the time of inflection*.

Another key question is when approximately that maximum is expected to occur. For that task we recall again the properties of *Gompertz* distribution and compute the value of the function at e times the inflection point

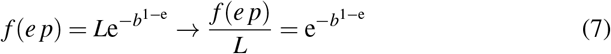

The above function converges to unity very rapidly for values of *b* > 1, so we have proven next Lemma 3.2.

#### Lemma 3.2.

*Given the assumptions of Lemma 3.1 the value of the cumulative variable under consideration at time e p is approaching fast the limit value for the current cycle of the disease*.

### 3.2. The Four Stages of a Disease in a Country

Although we can use the confirmed cases of a disease in order to study the time evolution, it is preferable to use the cumulative deaths because its count is more reliable, at least for countries that present a higher level of transparency, we can simply argue that “*dead cannot be hidden*”. So we focus on the curve of total losses until the current date, starting from the time of first reported loss in each country/region of interest.

#### 3.2.1. EDE approach

A first task is to use method *Extremum Distance Estimator* (*EDE*), see [5], [7], which is a rigid method in the sense that the inflection point has to be reached for a sufficient time in order to be able to give an estimation. The two critical points correspond to the two *Unit Invariant Knee* (*UIK*) points, see [6], for the first (convex) and the second (concave) part of the curve. Since curve begins its main exponential growth at the first *UIK* while after the relevant second enters in the slowing phase, we have an objective way to divide the process. If we follow the definitions of method *EDE*, then the sequence of points in ascending order is {*x*_1_, *χ*_*F*1_, *χ*_*D*_, *χ*_*F*2_, *x*_*n*_}, with *χ*_*D*_ the estimation for the inflection point, thus we can divide the time evolution in next four stages{*I* = [*x*_1_, *χ*_*F*1_), *II* = [*χ*_*F*1_, *χ*_*D*_), *III* = [*χ*_*D*_, *χ*_*F*2_), *IV* = [*χ*_*F*2_, *x*_*n*_]}.

In order to be able to infer about the percentage of completion for stage IV, given a death curve for a disease, we introduce next Definitions.

##### Definition 3.1.

*Let a partially or fully completed sigmoid curve for the cumulative deaths of a disease. Then if L*_*D*_ *is the observed losses at the inflection time t*_*i*_ *estimated by EDE and if L*_*max*_ *is the currently existing losses, the* Loss to Inflection Ratio (*LIR*_*D*_) *is*

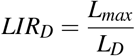

##### Definition 3.2.

*Let a given sigmoid curve for the cumulative deaths of a disease. Then if t is the current time in days from the first death and if t*_*D*_ *is the inflection time estimated by EDE, then* Time to Inflection Ratio (*T IR*_*D*_) *is*

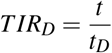

One of the most completed cycles in COVID–19 is the case of Austria, from the first death to May–27, see Fig. 4. There exist countries which have not reached their *EDE* inflection point and continue the stage II, see Fig. 5 for Brazil.

**Figure 4:**
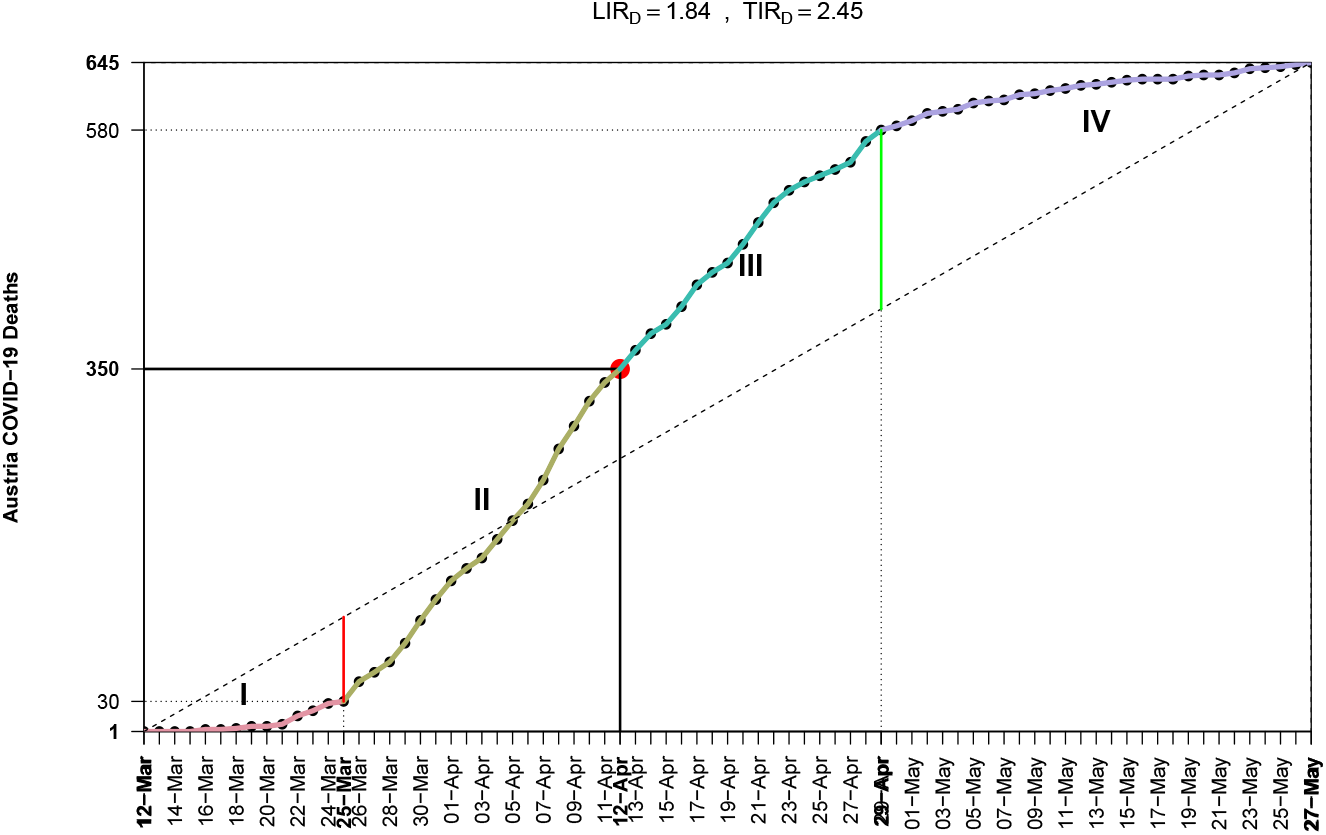
The 4 EDE-stages for COVID–19 in Austria

**Figure 5:**
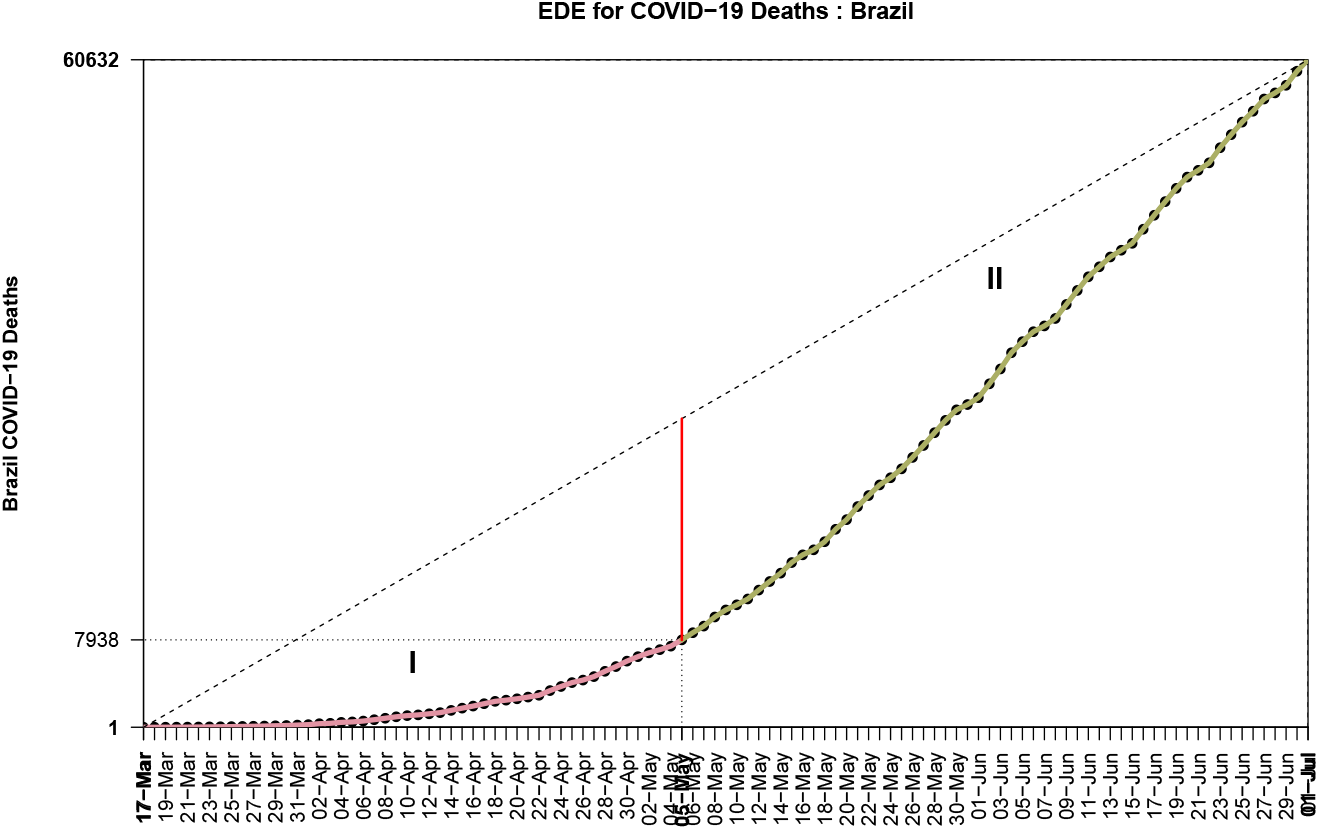
The first 2 EDE-stages for COVID–19 in Brazil

The *LIR*_*D*_ & *TIR*_*d*_ for the first 20 countries in the global list of deaths due to COVID–19 has been computed and is presented in Table 2^1^, which carries much information:

**Table 1:** *LIR*_*D*_, *TIR*_*D*_ for the first 20 countries in COVID–19 losses and the World (2020/07/02)

| Country | Begin | End | Inflection | Deaths | LIR | TIR |
| --- | --- | --- | --- | --- | --- | --- |
| US | 29-Feb | 01-Jun | 55810 | 106136 | 1.90 | 1.63 |
| Brazil | 17-Mar | 01-Jul |  | 60632 |  |  |
| United Kingdom | 06-Mar | 01-Jul | 21840 | 43991 | 2.01 | 2.44 |
| Italy | 21-Feb | 01-Jul | 17669 | 34788 | 1.97 | 2.79 |
| France | 15-Feb | 01-Jul | 15731 | 29864 | 1.90 | 2.32 |
| Mexico | 19-Mar | 23-May |  | 7179 |  |  |
| Spain | 03-Mar | 01-Jul | 16606 | 28364 | 1.71 | 3.08 |
| India | 11-Mar | 01-Jul |  | 17834 |  |  |
| Iran | 19-Feb | 20-May | 3294 | 7183 | 2.18 | 2.07 |
| Peru | 20-Mar | 01-Jul |  | 9860 |  |  |
| Belgium | 11-Mar | 01-Jul | 5683 | 9754 | 1.72 | 2.87 |
| Russia | 19-Mar | 01-Jul |  | 9521 |  |  |
| Germany | 09-Mar | 01-Jul | 4352 | 8995 | 2.07 | 2.92 |
| Canada | 09-Mar | 01-Jul | 4823 | 8678 | 1.80 | 1.87 |
| Netherlands | 06-Mar | 01-Jul | 3327 | 6132 | 1.84 | 2.85 |
| Chile | 22-Mar | 01-Jul |  | 5753 |  |  |
| Sweden | 11-Mar | 01-Jul | 2669 | 5370 | 2.01 | 2.15 |
| Turkey | 17-Mar | 01-Jul | 2376 | 5150 | 2.17 | 2.94 |
| China | 23-Jan | 07-Apr | 1369 | 3335 | 2.44 | 3.57 |
| Ecuador | 14-Mar | 01-Jul | 2145 | 4576 | 2.13 | 1.88 |
| World | 23-Jan | 27-May |  | 356707 |  |  |

**Table 2:**
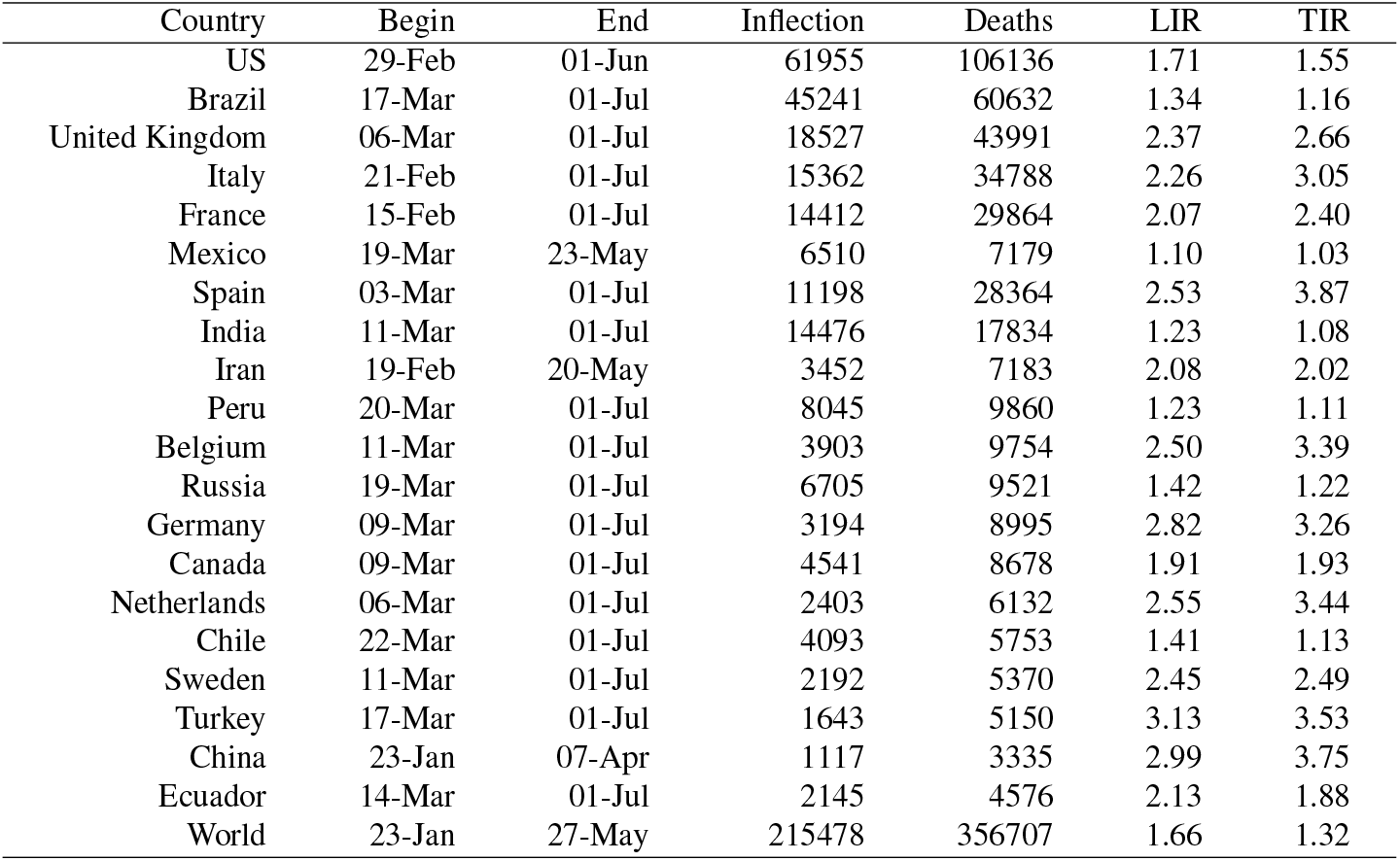
*LIR_S_, TIR_S_* for the first 20 countries in COVID–19 losses and the World (2020/07/02)

- First of all the fact that method *EDE* computes the inflection point as the midpoint between the lower and upper *UIK* point, guarantees that a value close to 2 is an index of completing a symmetric sigmoid curve. However not all diseases offer such an option, for example COVID–19 has shown highly non symmetrical behavior for all countries that completed the first cycle. China til April 14 (before changing the number of deaths) had a value of *LIR*_*D,China*_ = 2.44.
- If we take the observed values of *LIR*_*D*_ as proxies, then we can estimate a lower threshold for the expected losses from disease by simply doubling the *L*_*D*_.
- For countries that inflection point has not yet been reached, the expected losses will be more than twice the currently reported ones.

All inflection computations have been performed by using [8].

#### 3.2.2. ESE approach

If we apply the *ESE* method for estimation the inflection point of the curve of total losses, then curve classification changes because points of interest are different in that method. Now, according to the definitions of [5, 7], we focus on the x–left (*x*_*l*_) and x–right (*x*_*r*_) points where the left and right chord respectively are tangent to the curve. Of course here we use the estimated points *χ*_*l*_, *χ*_*r*_ due to the noisy nature of the curve. The sequence of points in ascending order is{*x*_1_, *χ*_*r*_, *χ*_*S*_, *χ*_*l*_, *x*_*n*_}, with *χ*_*S*_ the estimation for the inflection point, thus we can divide the time evolution in next four stages {*I* = [*x*_1_, *χ*_*r*_), *II* = [*χ*_*r*_, *χ*_*S*_), *III* = [*χ*_*S*_, *χ*_*l*_), *IV* = [*χ*_*l*_, *x*_*n*_]}.

Given the fact that the two methods *EDE, ESE* give different estimations, we need next Definitions

##### Definition 3.3.

*Let a partially or fully completed sigmoid curve for the cumulative deaths of a disease. Then if L*_*S*_ *is the observed losses at the inflection time t*_*i*_ *estimated by ESE and if L*_*max*_ *is the currently existing losses, the* Loss to Inflection Ratio (*LIR*_*S*_)*is*

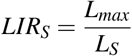

##### Definition 3.4.

*Let a given sigmoid curve for the cumulative deaths of a disease. Then if t is the current time in days from the first death and if t*_*S*_ *is the inflection time estimated by ESE, then* Time to Inflection Ratio (*T IR*_*S*_) *is*

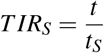

For Italy and Brazil we find Figs. 6, 7. Due to its design, *ESE* method can give an estimation for the inflection point at its early appearance and there is no need for a reasonable cycle completion to have been achieved. This is a reason for making it a good proxy for detecting the turning point of the disease from exponentially growing (Stages I, II) to slowing down (Stages III, IV). For example Brazil seems to have started the slowing down process.

**Figure 6:**
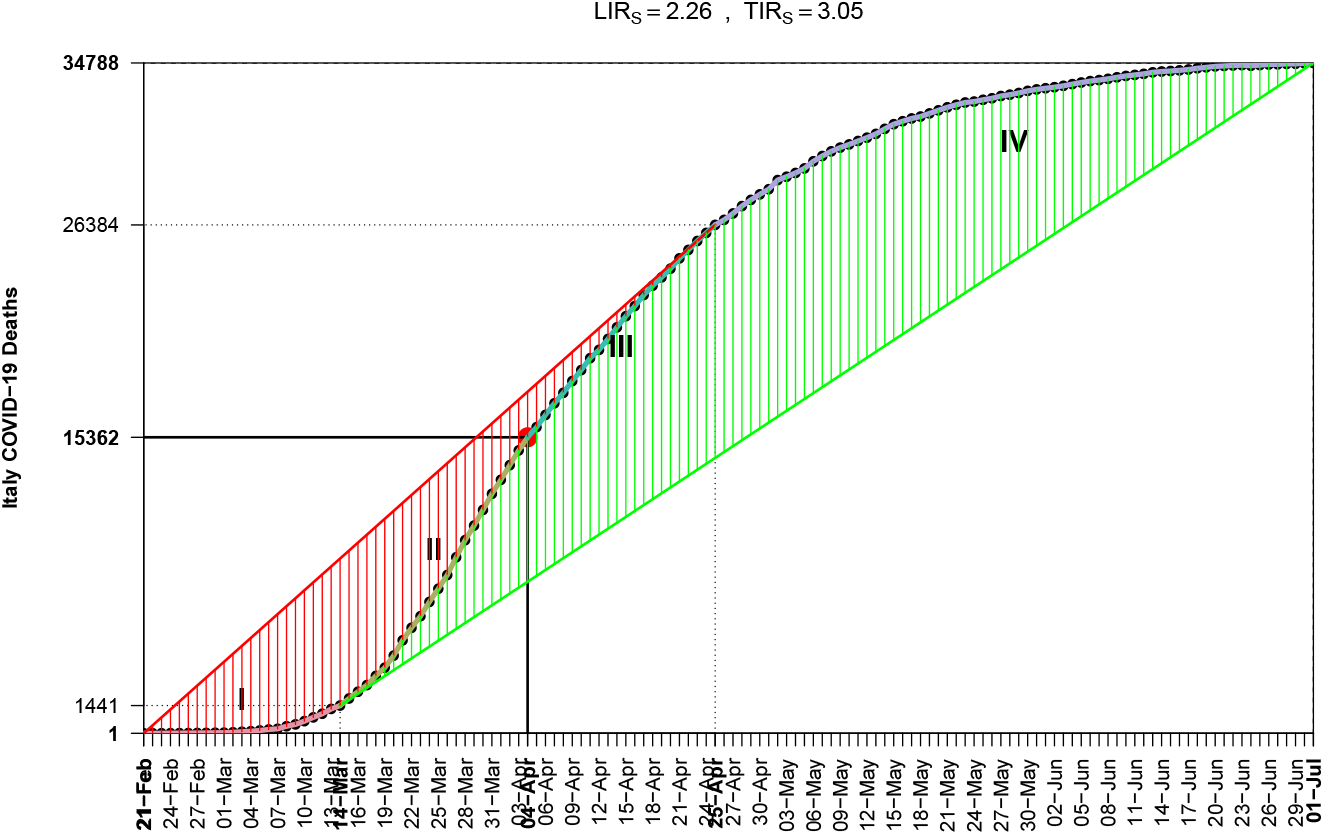
The 4 ESE-stages for COVID–19 in Italy

**Figure 7:**
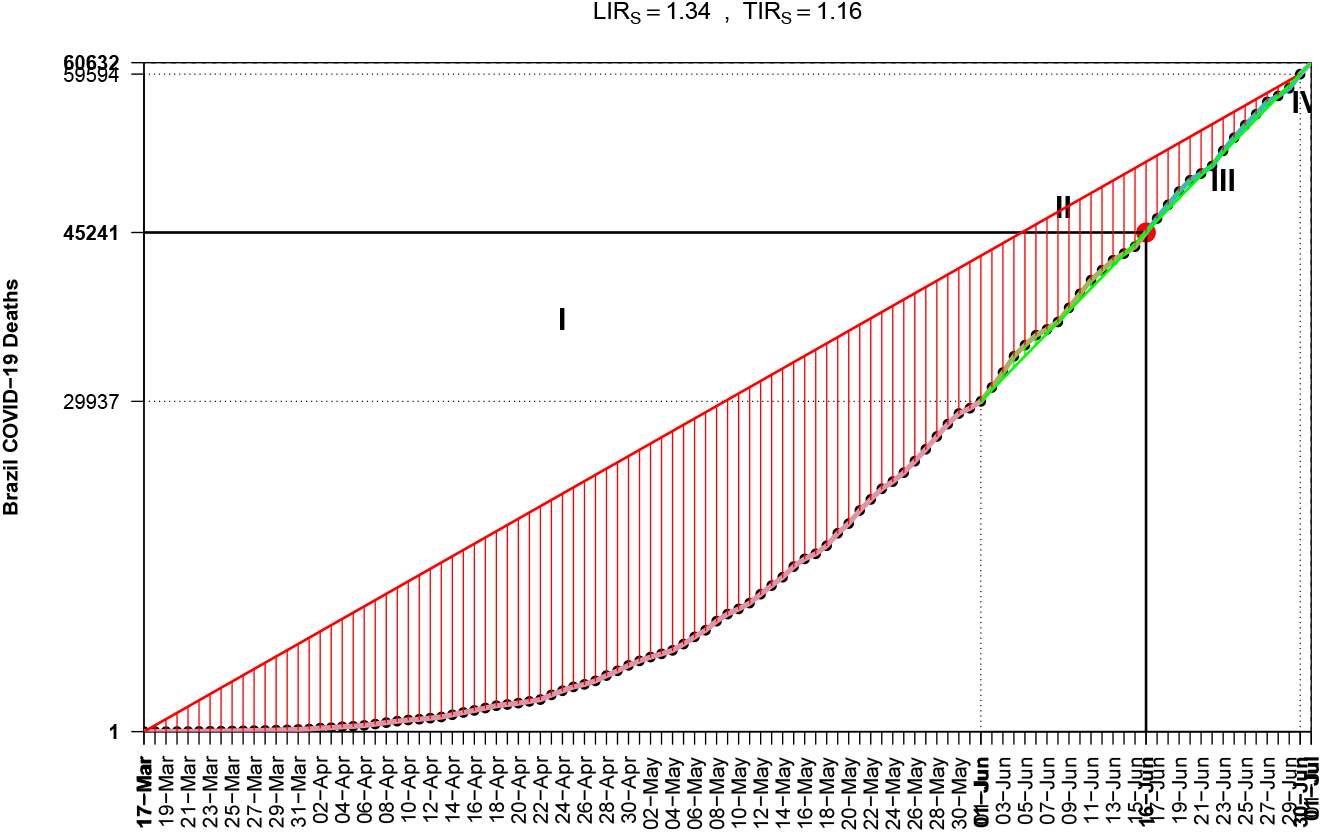
The not completed 4 ESE-stages for COVID–19 in Brazil

From Table 2, again created after using [8], we see that it is an encouraging sign the fact that al countries seem to have moved at an early version of Stage III.

### 3.3. Outbreak and Exponential Phase Times

It is interesting to mention that the time needed from the first reported case to the begin of the exponential phase, as measured by the first existing *UIK* point in the curve, varies from just 3 days (China, that’s an index that disease actually has began much before) to 94 days (India), see Fig. 8 for a graphic visualization of the countries with more than 50K confirmed cases, while in Table 3 they are listed in ascending order on the date of exponential begin – in its last lines there are mainly those from the Southern Hemisphere.

**Table 3:** Dates of first case and exponential growth begin for COVID–19 confirmed cases

| Country | Outbreak | Exponential | Days | Inflection |
| --- | --- | --- | --- | --- |
| China | Jan-22 | Jan-25 | 3 | Feb-09 |
| Italy | Jan-30 | Mar-10 | 40 | Apr-03 |
| Spain | Jan-31 | Mar-12 | 41 | Apr-05 |
| Germany | Jan-26 | Mar-12 | 46 | Apr-05 |
| France | Jan-23 | Mar-15 | 52 | Apr-06 |
| Netherlands | Feb-26 | Mar-17 | 20 | Apr-07 |
| Belgium | Feb-03 | Mar-19 | 45 | Apr-12 |
| Iran | Feb-18 | Mar-23 | 34 |  |
| US | Jan-22 | Mar-25 | 63 |  |
| United Kingdom | Jan-30 | Mar-25 | 55 | Apr-24 |
| Canada | Jan-25 | Mar-25 | 60 | Apr-26 |
| Turkey | Mar-10 | Mar-26 | 16 | Apr-15 |
| World | Jan-22 | Mar-30 | 68 |  |
| Sweden | Jan-30 | Mar-31 | 61 |  |
| Ecuador | Feb-29 | Apr-07 | 38 | Apr-29 |
| Belarus | Feb-27 | Apr-14 | 47 | May-18 |
| Russia | Jan-30 | Apr-16 | 77 | May-23 |
| Qatar | Feb-28 | Apr-21 | 53 | May-28 |
| Peru | Mar-05 | Apr-24 | 50 | Jun-01 |
| Saudi Arabia | Mar-01 | Apr-25 | 55 |  |
| Indonesia | Mar-01 | May-03 | 63 |  |
| Mexico | Feb-27 | May-05 | 68 |  |
| Pakistan | Feb-24 | May-06 | 72 |  |
| India | Jan-29 | May-09 | 101 |  |
| Bangladesh | Mar-07 | May-10 | 64 |  |
| Colombia | Mar-05 | May-11 | 67 |  |
| Brazil | Feb-25 | May-12 | 77 |  |
| Chile | Mar-02 | May-12 | 71 | Jun-13 |
| Egypt | Feb-13 | May-15 | 92 |  |
| Argentina | Mar-02 | May-18 | 77 |  |
| South Africa | Mar-04 | May-20 | 77 |  |
| Iraq | Feb-23 | May-27 | 94 |  |

**Figure 8:**
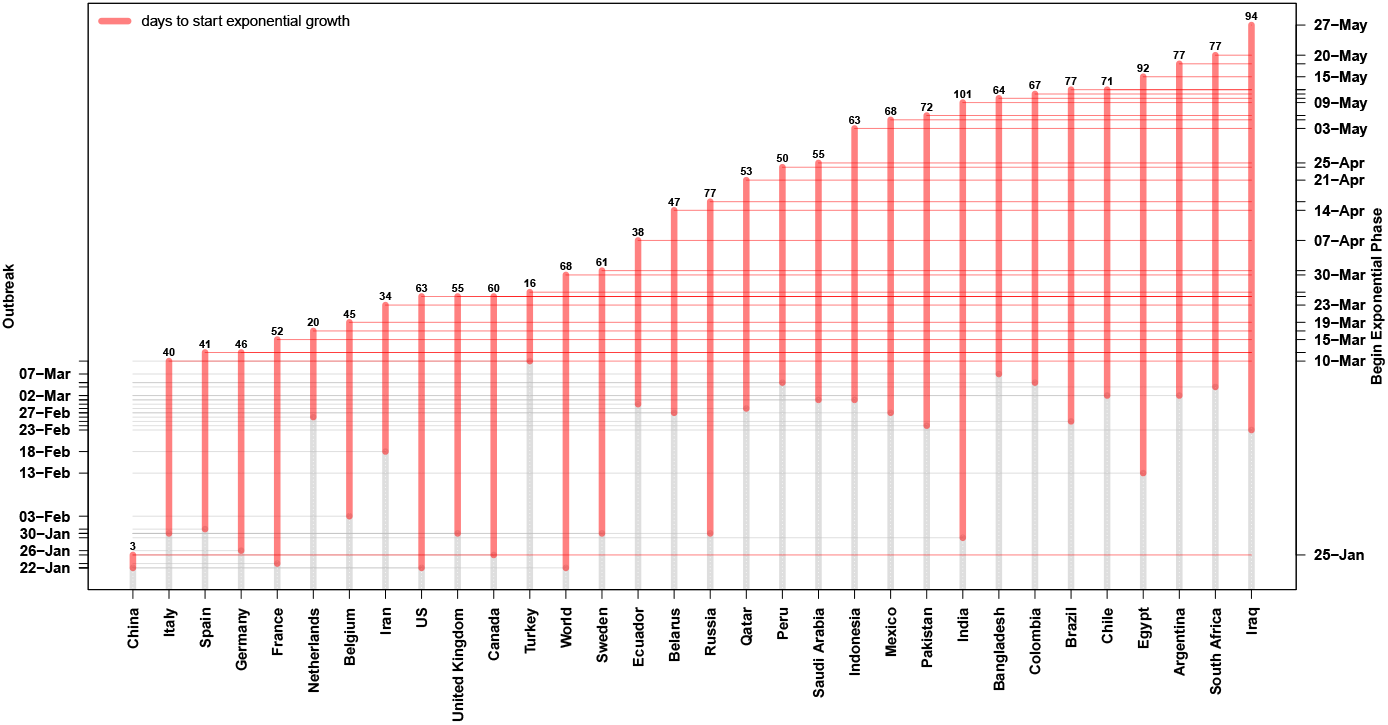
Time to start the exponential phase after the COVID–19 outbreak (confirmed cases)

### 3.4. The Time Dynamics of Inflection Point as Percentile Point

After having a reasonable number of daily data we area ble to compute the Inverse Empirical Cumulative Distribution Function (IECDF) for the variable of interest, usually for deaths from the disease. That curve becomes a sigmoid one after the overall cumulative process has reached its inflection point and we can estimate it either by *EDE* or *ESE* method. That point is a percentile one for the empirical distribution and has always a certain movement from a value close to unity, say 0.95 just after inflection, to a value slightly over 0.10 at the end of current disease cycle.

For the 20 most affected countries plus World, the relevant plots for EDE (past month) and ESE(past 2 months) can be found at Figs. 9 and 10. Obviously it is evident a country classification according to its inflection dynamics, since IECDF is comparable between them. There are countries like China, Spain, Italy which seem to have completed their first cycle. Other countries like UK, France tend to reach that task now. A third class is countries like US, Canada, Ecuador that are at the last miles. Finally a fourth class contains Russia, Brazil, Peru, India that are still at the first exponential phase, actually they have not reached yet an *EDE* inflection point. From the plots of 20 most affected countries we can derive two next

**Figure 9:**
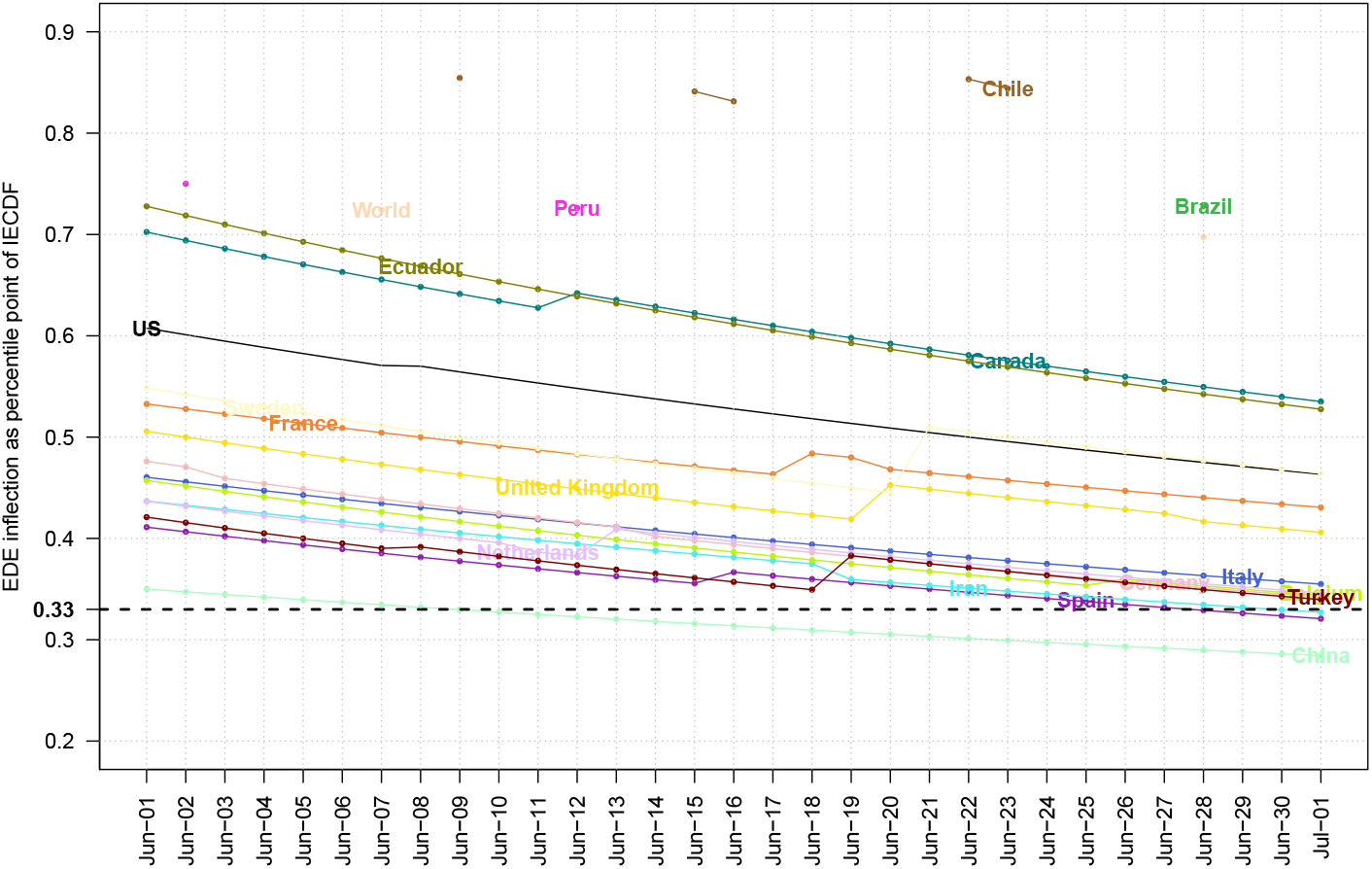
Time evolution of *EDE* inflection as percentile for most affected by COVID–19 countries

**Figure 10:**
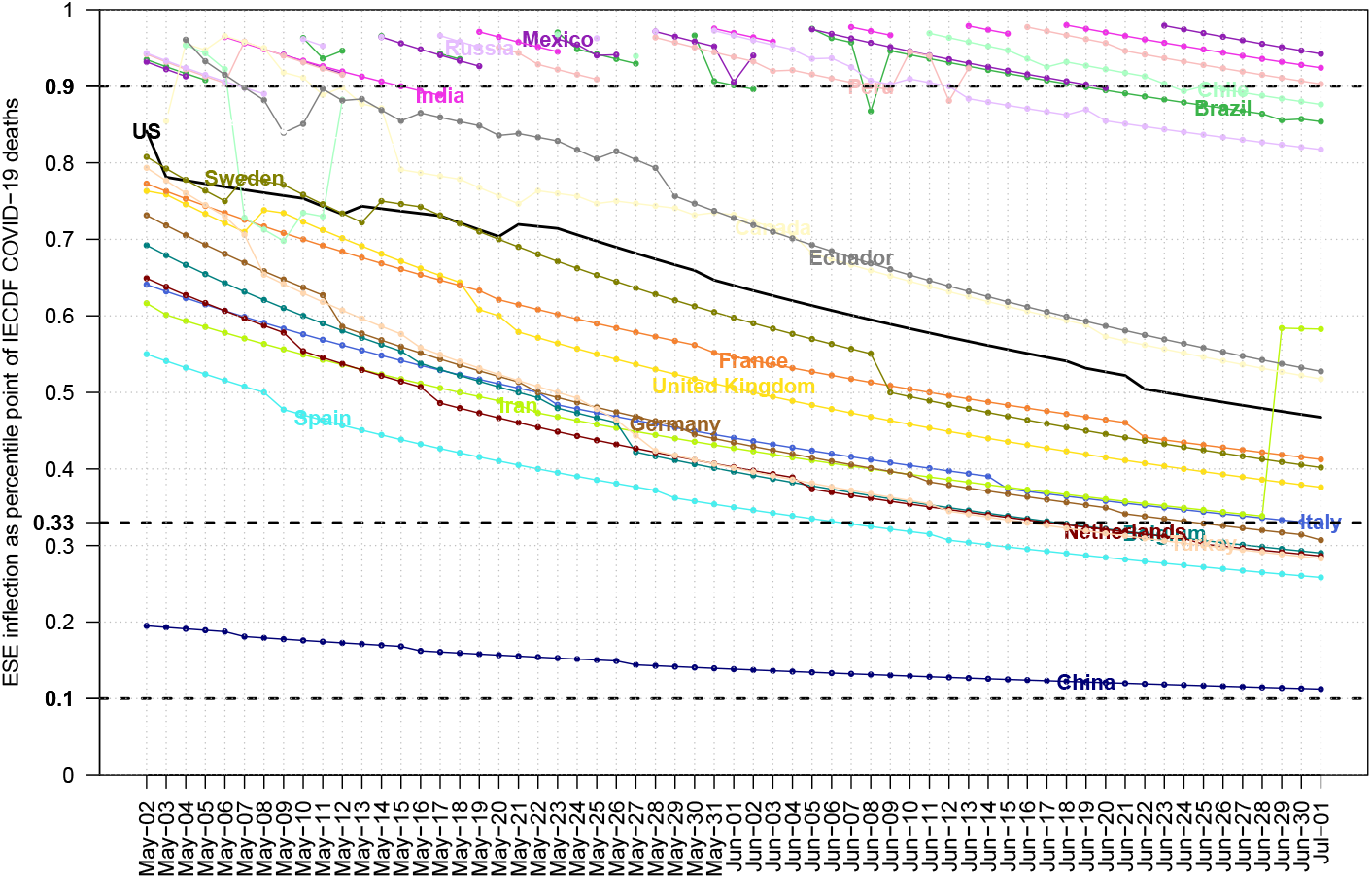
Time evolution of *ESE* inflection as percentile for most affected by COVID–19 countries

#### Empirical Observation 3.1.

*The inflection point of IECDF curve which is computed by EDE needs substantial time to be reached and then tends towards the 1/3 percentile point after a few months*.

#### Empirical Observation 3.2.

*The inflection point of IECDF curve which is computed by ESE although it can be found at an earlier stage, then it needs more time for converging towards either to the 1/3 or 1/10 percentile point*.

## 4. Comparison of Approaches

Since the COVID–19 has completed its first cycle in many countries and has also just begun for others, we are able now to compare the two approaches, the broadly accepted mathematical models (*SEIR* etc) and the novel suggested by this work. That task is better to be done for UK, US and Greece because they are representative cases for a big, medium and small country respectively, while there exist works published online about them.

### 4.1. UK

The first approach is the mathematical model used by [11] with data til March 16th. Their prediction for UK was presented at Fig. 1 which is a totally symmetric curve. The model output had a cumulative value of 510K for losses in UK, far beyond the currently reported 44K.

The alternative approach, based on data available til March 16th, gave that UK was just on the exponential phase and no inference could be done at that early time. However, after passing a reasonable time period, method *ESE* first and then *EDE* gave their estimations, see Fig. 11. We observe that middle May, both methods converged to the value of 18527, thus an estimation for the total losses in UK begins with that number as multiplier and given China’s *LIR*_*S*_ = 3, we may expect at the worst cases approximately 55K losses, again much less than the 510K, actually close to the 10% of model’s predicted value. From the sample presented at Fig. 11 we were able to extract the inflection point at May 24th which has a value of 255K due to the symmetry of the curves, while the robust estimation of 18257 was found by both methods at May 9th.

**Figure 11:**
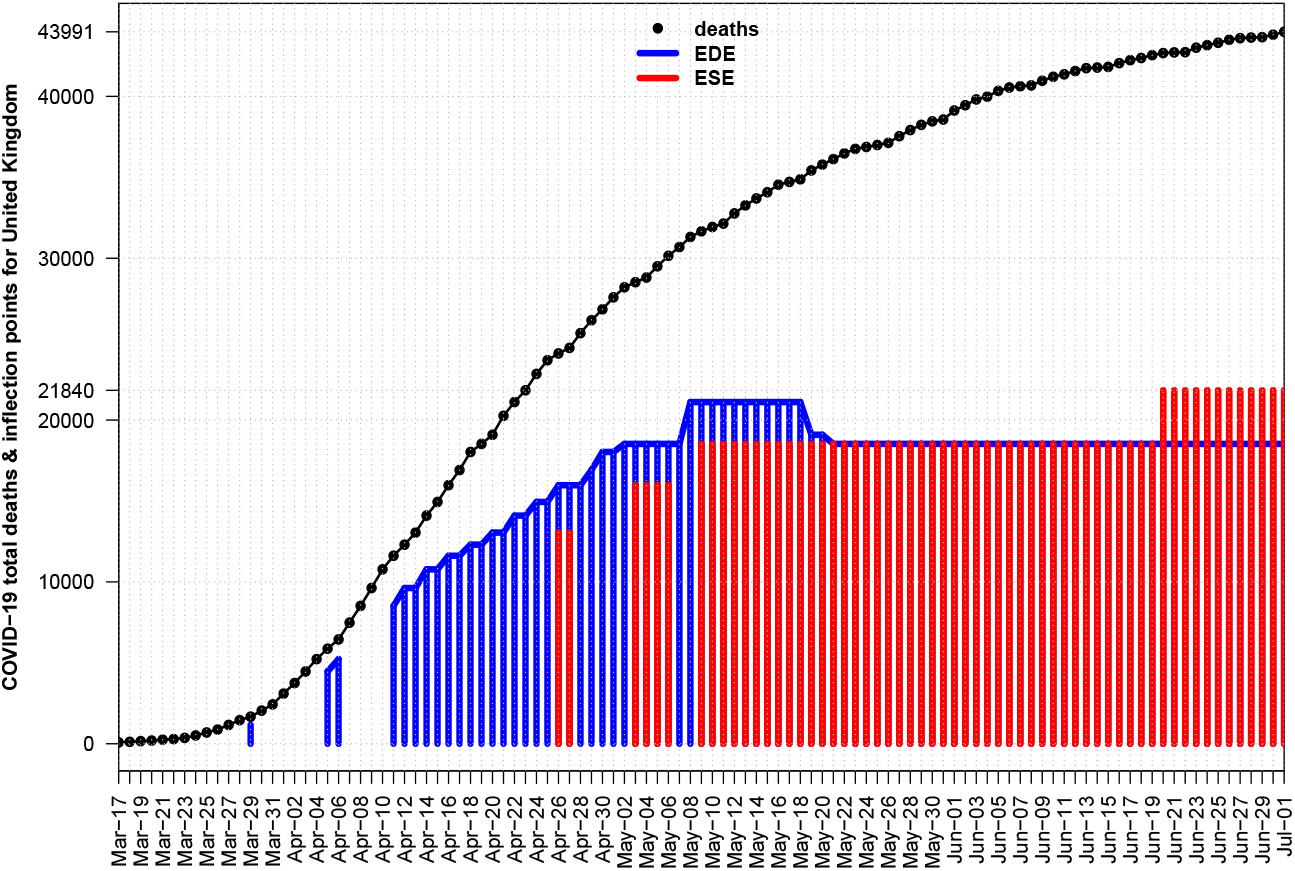
History of EDE & ESE inflection estimations for UK

### 4.2. US

From the same Fig. 1 of [11] we find that expected evolution by mathematical models is of symmetric shape and predicts 2.2M losses for US after using data available til March 16th. Our alternative approach again would not give any kind of estimation til March 16th, since the disease was at Stages I, II, but it would be able to create *ESE* estimations for inflection point at early April, while then it had an increasing trend. *EDE* method which is more rigid began to give output at early May and both methods have finally converged to the values of 61252(2020-04-29), 55162(2020-04-26) respectively given order of mentioning above, see Fig. 12. Thus if we apply the ‘*e-rule*’ here or if we take the worst case scenario, i.e. if we use China’s *LIR*_*S*_ = 3 as multiplier, then we estimate [167K,184K] (*ESE*) and [150K,165K] (*EDE*), again less than the 10% of predicted 2.2M.

**Figure 12:**
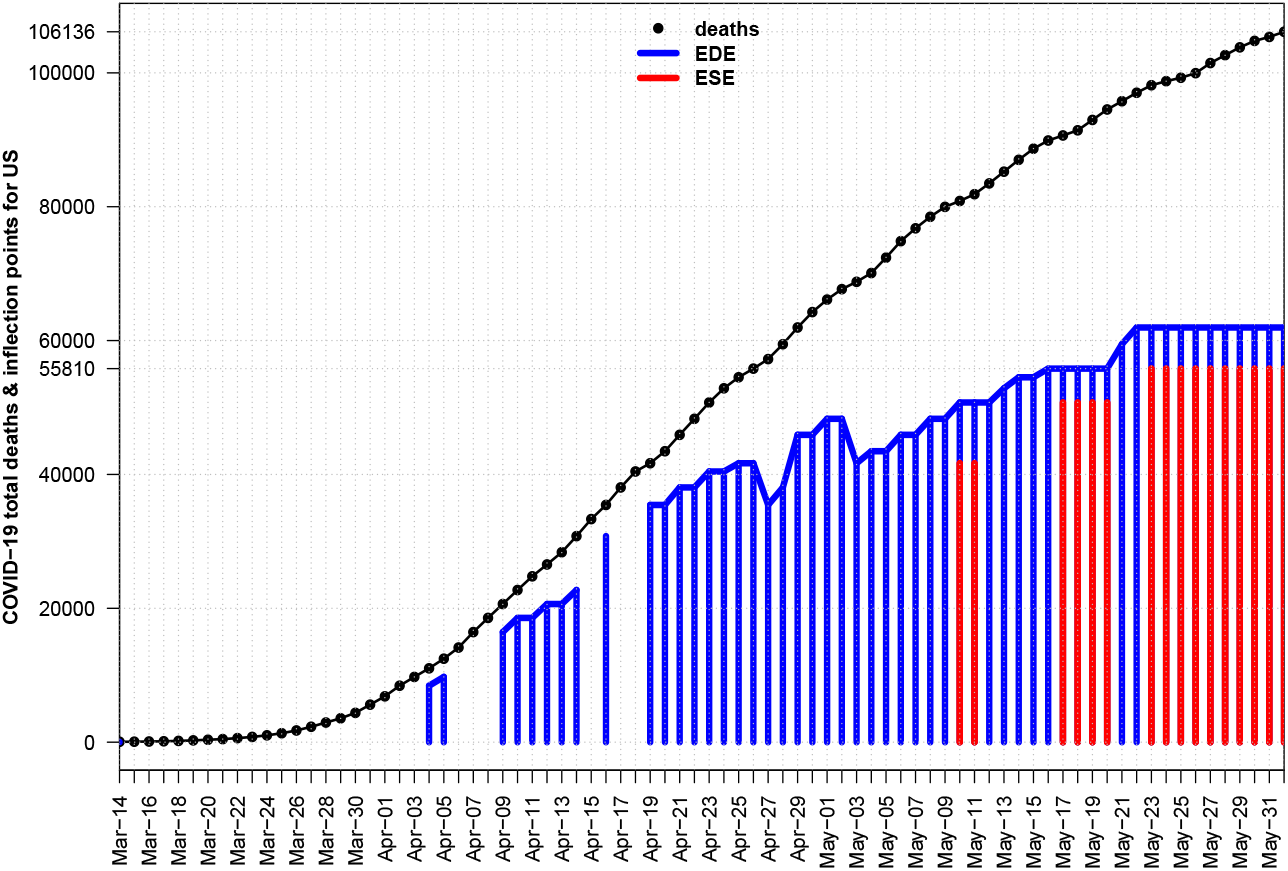
History of *EDE* & *ESE* inflection estimations for US

### 4.3. Greece

The model predictions of [21] are presented at their Fig. 1 where the model has been fitted to the available data til April 26. Although non published, the estimation given by the senior author of [21] for the deaths expected in Greece if no measures had been taken was a number of 13685 losses (as stated in the last daily TV briefing of May 26th). We may probably compare that number with Belgium’s 9334 deaths for the same date, however Greece is not such an extroverted country (recall that Belgium is the EE administrative center and many other International organisations are based in Belgium), so given that Greece has 1 million less population, that estimation would not have been realized.

If we had applied our alternative view for the same time period, then we could had created Fig. 13 and on April 26th we could had made a prediction for the total deaths by using the ‘*e-rule*’ finding for the worst case, based on China, a number of 171 deaths, very close to the finally obtained from that cycle in Greece (about May 24th).

**Figure 13:**
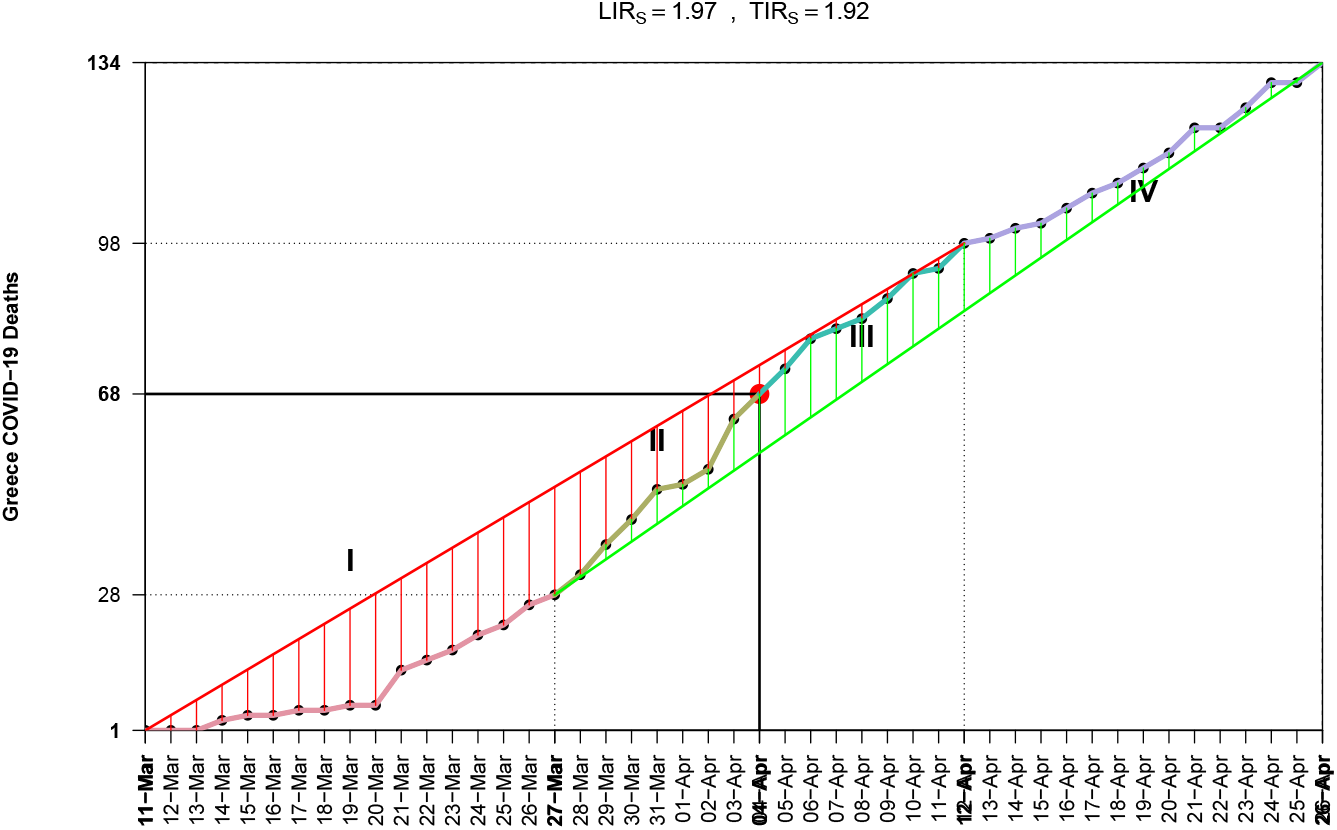
The 4 EDE-stages for COVID–19 in Greece for the study time of [21]

## 5. World Dynamics: Number of Expected Cycles and Losses

The fact that Earth is divided in Northern and Southern Hemispheres is proven to play a critical role for the number of COVID–19 cycles. First it was the winter at North and that gave us the first cycle. Now it is the winter at South and a new cycle has began. We can estimate the approximate begin date if we apply method *EDE* for the curve of global cumulative deaths, after its inflection point. Then we find a new turning point on May–27, see Fig. 14, where we have also plotted the *LOWESS* smooth curve, after [9]. It is certainly not a good sign which shows that disease will not slow down unless it will have covered both winters on Earth. Except the geographical reasoning, we have observed the appearance of a second cycle in countries like Iran which after May 19th has increasing number of deaths again.

**Figure 14:**
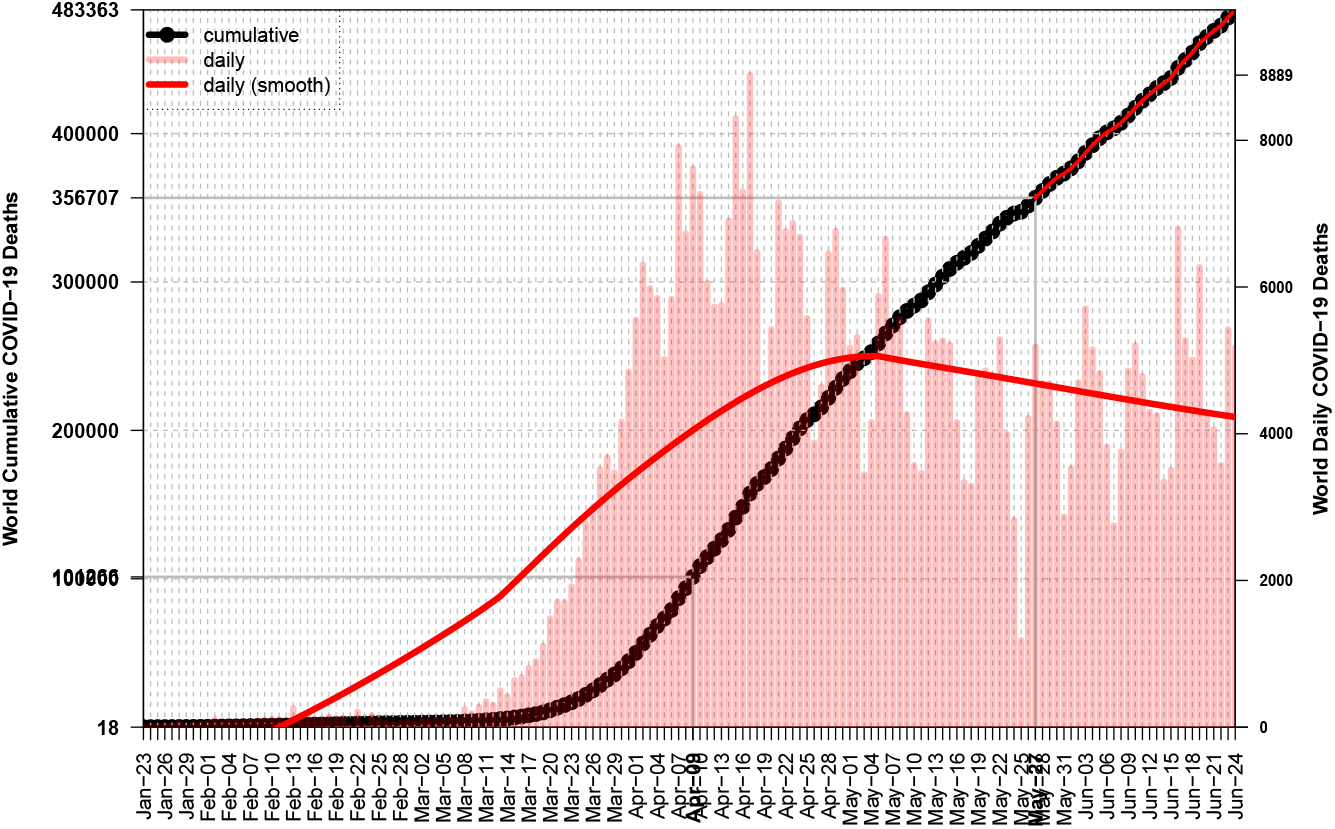
World has began a new COVID–19 Deaths cycle on May–27

We have seen from Tables 1 & 2 that World has not reached an inflection point under method *EDE* for the first cycle, while the relevant point that was found by method *ESE* gave rather small values for *LIR*_*S*_ = 1.66 & *TIR*_*S*_ = 1.32, thus we cannot make any robust inferences for the expected number of losses. If we accept that second cycle will have the same intensity as the first, then we may expect a slightly less number than 356K of May–27, as extra deaths. If we also accept the hypothesis that in Northern Hemisphere only Russia has not completed its cycle, then we may not expect many losses from there.

Unfortunately one of the most powerful countries of the World, United States, seem to have began a second cycle after June 1st, see Fig. 15.

**Figure 15:**
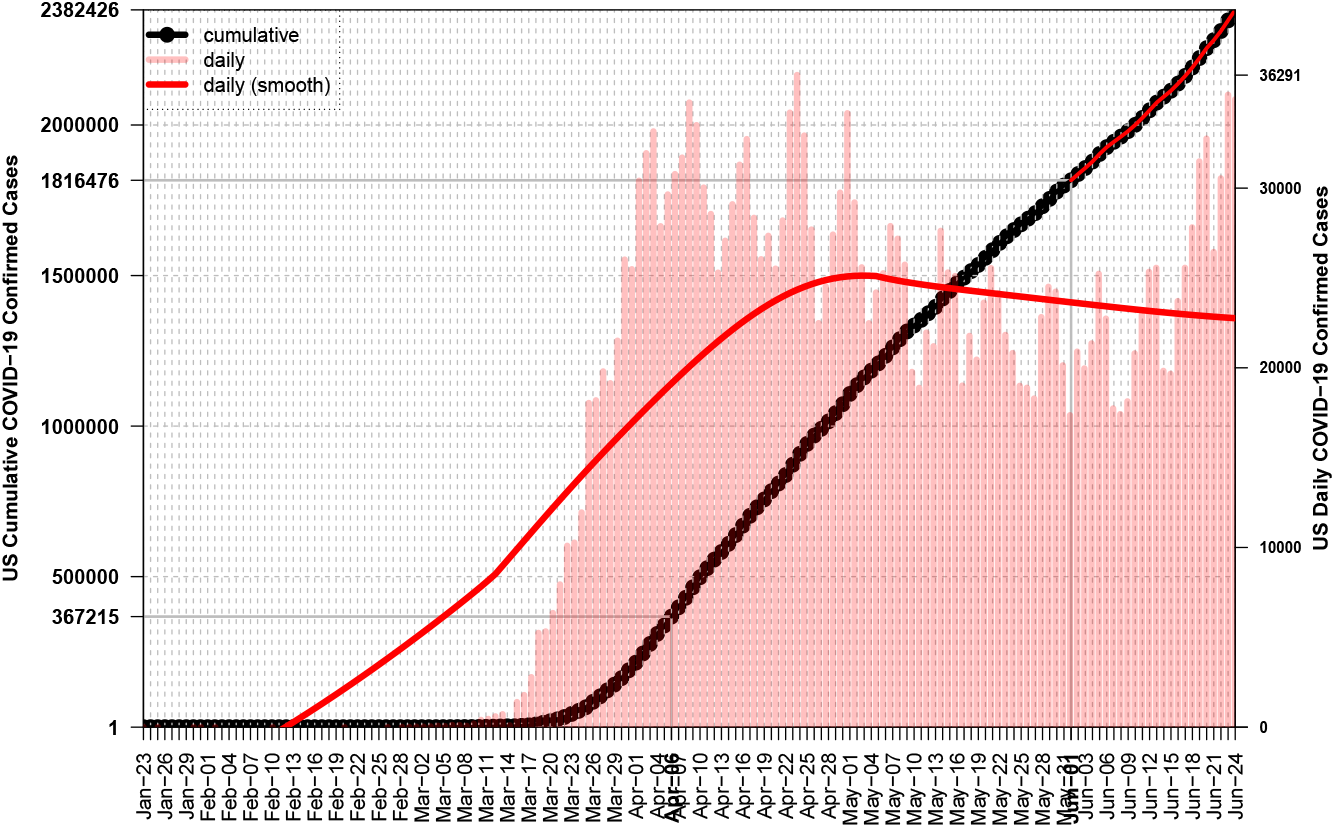
US daily confirmed cases for COVID–19 began to increase again after June–1

## 6. Discussion

As usually, since truth is somewhere in the middle, we cannot reject the currently accepted set of mathematical models and the underlying treatment of a disease, but we cannot rely only on them for estimating the time evolution of the pandemic. Although the effective reproduction rates *R*_0_, *R*_*t*_ can serve as reliable proxies of the picture taken at the outbreak or later, the recent COVID–19 adventure has shown that the requirement of *R*_*t*_ *<* 1 has a very limited usefulness for either predicting the outcome of a disease cycle or for controlling the pandemic. Examples are almost all affected Western countries, where the above condition although fulfilled, both confirmed cases and deaths did not slow down until they had created a sigmoid curve for the relevant cumulative variables. Coronavirus seems to act like a ‘*disease weapon bomb*’ which after being released has an autonomous route despite the *NPIs* that a country will apply or not.

The successful cases for combating the virus attach have been only those countries (like Greece) or territories (like Zahara de la Sierra of Spain) that applied a very strict policy of almost total isolation from anything that was arrival from abroad. Recall that in Greece at the very early stage, neither the Sailing Yachts were allowed to moor at the ports. That strict isolation combined by the national lockdown and given the country terrain, led to the almost zero deaths compared to other affected countries. Especially for Greece next particularities hold:

- Greece presents difficulties in communication between its provinces due to the terrain geography (mountains, many small islands)
- The prohibition of airport flights from abroad reduced the import of new cases, something that other countries were forced to do much later than Greece
- The non existence of a direct flight from Wuhan did not transmit the virus directly from its first place of appearance, unlike Italy which had a stronger connection to China

The specific country constraints should have been considered for a less hard lockdown in the Aegean Islands, since isolation was almost absolute due to the lack of transports. For example in a mountain village with population 280, what was the rational of not permitting local residents to walk free of constraints? Who was going to infect whom? Those measures were extremely hard for the already social self isolated villages and towns in Greece and did not offer any kind of help against the virus. Thus decision making based merely on the values of *R*_*t*_ could be just harmful for the society, it can act as a “bulldozer in the garden”.

Since cumulative deaths from any disease follow an S-shape or sigmoid curve, then it is easy to compute the inflection point by using methods *EDE* & *ESE* and then define new concepts for helping us to monitor the time evolution and to compare different countries. That is the contribution of current work and we hope subsequent works to improve the newly introduced concepts.

## Data Availability

COVID-19 Data Repository by the Center for Systems Science and Engineering (CSSE) at Johns Hopkins University

https://github.com/CSSEGISandData/COVID-19

## Footnotes

1 Til the end of first cycle: US June–1, Spain May–21, Iran May–19, China Apr–6, World May–27

